# Identifying barriers to genetic testing in subspecialty cardiac care

**DOI:** 10.1101/2024.05.15.24307347

**Authors:** Sierra Pond, Genevie Echols, Martin M Tristani-Firouzi, Susan P Etheridge, Hannah S Anderson, Briana L Sawyer

## Abstract

**Background:** The utility of genetic testing in cardiovascular medicine is well-established in expert consensus statements for optimizing patient care. However, significant genetic testing care gaps persist for patients with inherited cardiovascular conditions.

**Objective:** This study aimed to understand why genetic testing care gaps in cardiovascular medicine exist by evaluating cardiovascular providers’ opinions and use of genetic testing.

**Methods:** We developed and administered an anonymous survey to cardiovascular providers delivering direct patient care in the United States. Participants were contacted in collaboration with the Sudden Arrhythmia Death Syndromes (SADS) Foundation.

**Results:** A total of 111 individuals completed the survey representing the following specialties: electrophysiology (55%, n=61), general cardiology (10.8%, n=12), imaging (7.2%, n=8), heart failure/transplant (6.3%, n=7), interventional cardiology (6.3%, n=7), fetal cardiology (5.4%, n=6), and other (9%, n=10). Eighty-six percent of respondents (n=97) stated genetic testing is ‘very relevant’ in the care of their patients. Eighty percent of electrophysiologists reported ordering genetic testing a few times a month or more. Navigating insurance authorization and billing procedures was an identified area of discomfort by 47.7% of respondents (n=53). Overall, cardiovascular provider specialty was shown to impact how often genetic testing is ordered. Provider work setting was shown to impact opinion of the utility of genetic testing for family screening, opinion of the utility of genetic testing for medication and device management, and how often genetic testing is ordered.

**Conclusions:** The results of this study support targeted provider education to increase the uptake of genetic testing for patients with inherited cardiovascular conditions.

## INTRODUCTION

Genetic testing as a diagnostic and management tool is a key component of patient care in the emergence of precision medicine. Inherited cardiac conditions are associated with substantial morbidity and mortality with known benefits from genetic testing to manage patients with inherited cardiac conditions [1-4]. Examples of the diagnostic utility of genetic testing in cardiovascular medicine include the ability to establish a molecular clinical diagnosis, earlier diagnosis, characterization of a patient’s syndrome subtype, elimination of other suspected conditions, and initiation of family cascade screening once a proband’s genetic variant has been identified [1, 3, 5-8]. The utility of genetic testing in cardiovascular management can be divided into two categories: prognostic and therapeutic. Examples of prognostic management enabled by genetic testing for cardiovascular patients include genotype-phenotype correlations [9] and anticipating patient prognosis with specific recommendations about physical activity and other lifestyle modifications such as drug avoidance [2, 10, 11]. Additionally, genetic testing facilitates therapeutic management decisions for cardiovascular patients regarding tailored pharmacological and procedural decisions [12, 13] and cardiac device management [12].

Guidelines and expert consensus statements supporting the use of genetic testing in cardiovascular conditions have been reported in the literature since 2011 [14, 15]. Since then, the specific language surrounding the use of genetic testing for patients with suspected cardiovascular conditions has evolved significantly [16] with new evidence to support and strengthen recommendations. Consensus statements and expert working groups on heritable cardiac conditions now encompass a wide range of diagnoses including familial hypercholesterolemia [17], adult congenital heart disease [18], hypertrophic cardiomyopathy [19], channelopathies [20], and arrhythmogenic cardiomyopathy [21].

The benefits of genetic testing in cardiology are not universally accepted and applied [22]. According to a 2023 statement from the American College of Cardiology, “patients with certain inherited cardiovascular conditions rarely receive genetic testing recommended by clinical practice guidelines” [23]. Genetic testing uptake is below 10% across all inherited cardiac conditions [23-25], which is in stark comparison to the strong recommendation of its utility. Previous research identified barriers to the use of genetic testing in other disciplines outside of cardiovascular medicine. Such barriers exist at the provider level, the clinic level, and the societal level at large, leaving genetic testing an ‘untapped resource’ [26] for patient care. Literature on barriers to genetic testing in other disciplines including cancer and neurology exists at length. Previous research on the barriers to the use of genetic testing in the world of subspecialty cardiac care focused on the patient perception of genetic testing [27] and practitioners’ confidence and desires for education in cardiovascular and sudden cardiac death genetics [28]. Past research has also demonstrated that there is no significant psychological harm done to patients who undergo cardiovascular genetic testing [29].

It remains unknown in cardiovascular medicine how provider specialty influences a provider’s opinion of, use of, and access to genetic testing. This project aims to assess the opinions of cardiovascular providers in various subspecialties on genetic testing; how cardiovascular providers implement genetic testing in their practice; and what barriers cardiovascular providers identify to their use of genetic testing. As a secondary aim, this project also aimed to compare these barriers among different provider types to assess differences between provider groups.

## METHODS

### Study Population

To assess the opinions of, use of, and barriers to the use of genetic testing in subspecialty cardiac care, a one-time quantitative survey was developed and administered to licensed cardiovascular providers who engage in direct patient care in the United States. To meet the inclusion criteria for survey participation, participants were required to self-identify as a physician, physician assistant, nurse, or advanced practice nurse (listed as MD, DO, PA, RN, or APRN in the survey). Genetic counselors and medical geneticists were intentionally excluded from participation in the survey to reduce bias.

### Survey Development

Questions were designed to elicit cardiovascular providers’ opinions of, use of, and access to genetic testing. Survey development was guided by previous research [28]. Our survey consisted of five main sections with twenty-nine total questions: (1) inclusion criteria, (2) provider opinions on genetic testing utility, (3) provider-reported implementation of genetic testing in their practice, (4) provider-identified barriers to the use of genetic testing in their practice, and (5) demographics. Survey questions were mainly Likert-scale and categorical, asking participants to define their opinion of genetic testing, their use of genetic testing, and their comfort with activities related to genetic testing. The survey was piloted by three pediatric cardiologists, three genetic counselors, and three members of the Sudden Arrhythmia Death Syndromes (SADS) Foundation before dissemination.

### Survey Administration

The anonymous electronic survey was administered via RedCap Software hosted at the University of Utah [30, 31]. The survey was fielded between January and April 2024. Participants provided written informed consent at the start of the survey. The study was approved and granted exemption status by the University of Utah’s Institutional Review Board (00164238).

Participants were contacted via email recruitment across the United States with integral help from the SADS Foundation. The recruitment email explained the intent of the research as well as the anonymous response platform. Providers from the SADS Foundation Physician Referral Network (PRN) were contacted first to establish contact with major healthcare institutions across the United States. Participants were encouraged to disseminate the survey within their professional networks. Professional societies were also targeted for contact including the Heart Failure Society of America and the Heart Rhythm Society. An intentional approach was taken to recruit participants from a representative group of cardiovascular subspecialties, patient age ranges, geographic regions, and a diverse range of workplace settings.

### Statistical Analysis

Means, standard deviations, and proportions were calculated for categorical variables. Chi-squared tests were used to compare categorical variables. Testing correction using the Bonferroni method was then performed as multiple testing comparisons were performed. The adjusted significance level was *P<0*.*0022*. Data analysis was performed with Python software version (3.9.5). Data are available upon request.

## RESULTS

### Demographics

A total of 125 individuals submitted responses to the survey. Twelve of these responses were excluded as they did not meet inclusion criteria or were incomplete. This resulted in 111 total responses for inclusion in the study analysis. The demographic characteristics of the study participants are summarized in Table 1. Most survey respondents (55%; n=61) reported working in the specialty of electrophysiology. Other major provider specialties included general cardiology (10.8%; n=12), imaging (7.2%; n=8), heart failure/transplant (6.3%; n=7), and interventional cardiology (6.3%; n=7). Most survey respondents (43.2%; n=48) reported seeing a mix of pediatric and adult cardiovascular patients. Thirty-two percent of respondents (n=36) reported seeing pediatric cardiovascular patients only; 22.5% of respondents (n=25) reported seeing adult cardiovascular patients only; and 1.8% of respondents (n=2) reported seeing prenatal cardiovascular patients. The geographic region and work setting of participants was primarily urban (82.9%; n=92) at university medical centers (78.4%, n=87). Most survey respondents (52.2%, n=58) reported having been in practice for more than fifteen years. Due to the wide variety of recruitment techniques, a survey response rate is not possible to calculate.

**TABLE 1.**
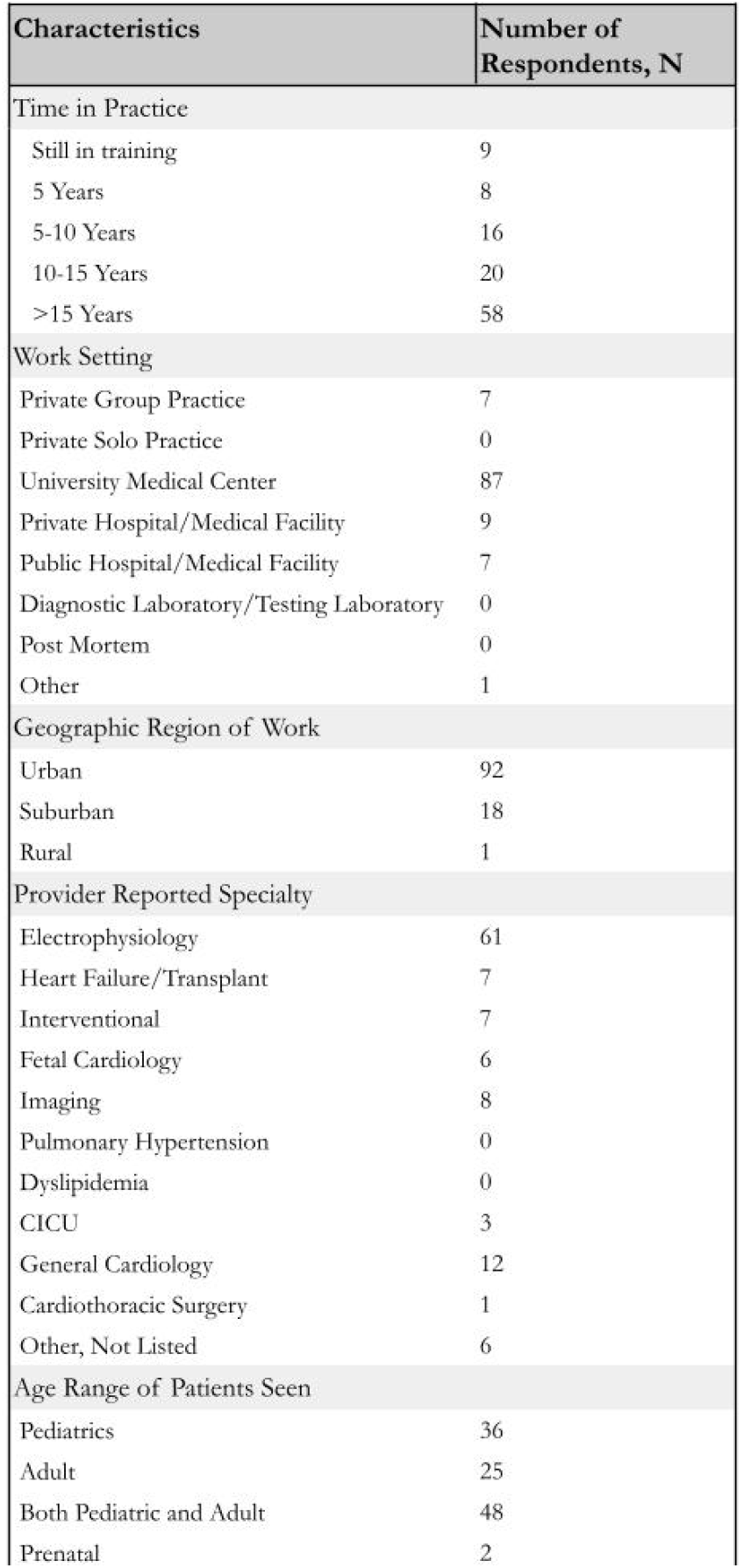
Demographic Characteristics of Study Participants. *Participants were asked a handful of demographic questions about their practice in section (5) of the survey. Data are reported in the number of respondents (n) to each question*.

### Opinion and Use of Genetic Testing

The majority of survey respondents (86%; n=97) reported genetic testing being ‘very relevant’ in the care of their patients. No survey respondents reported genetic testing being ‘not at all relevant in the care of their patients’. Figure 1 delineates responses to this question on the relevance of genetic testing in the care of patients by provider-reported specialty. Of those respondents who answered genetic testing is either ‘somewhat relevant’ (9.8%; n=11) or ‘slightly relevant’ (3.6%; n=3) in the care of their patients, provider specialties included electrophysiology, interventional cardiology, fetal cardiology, general cardiology, cardiac ICU, and imaging.

**FIGURE 1.**
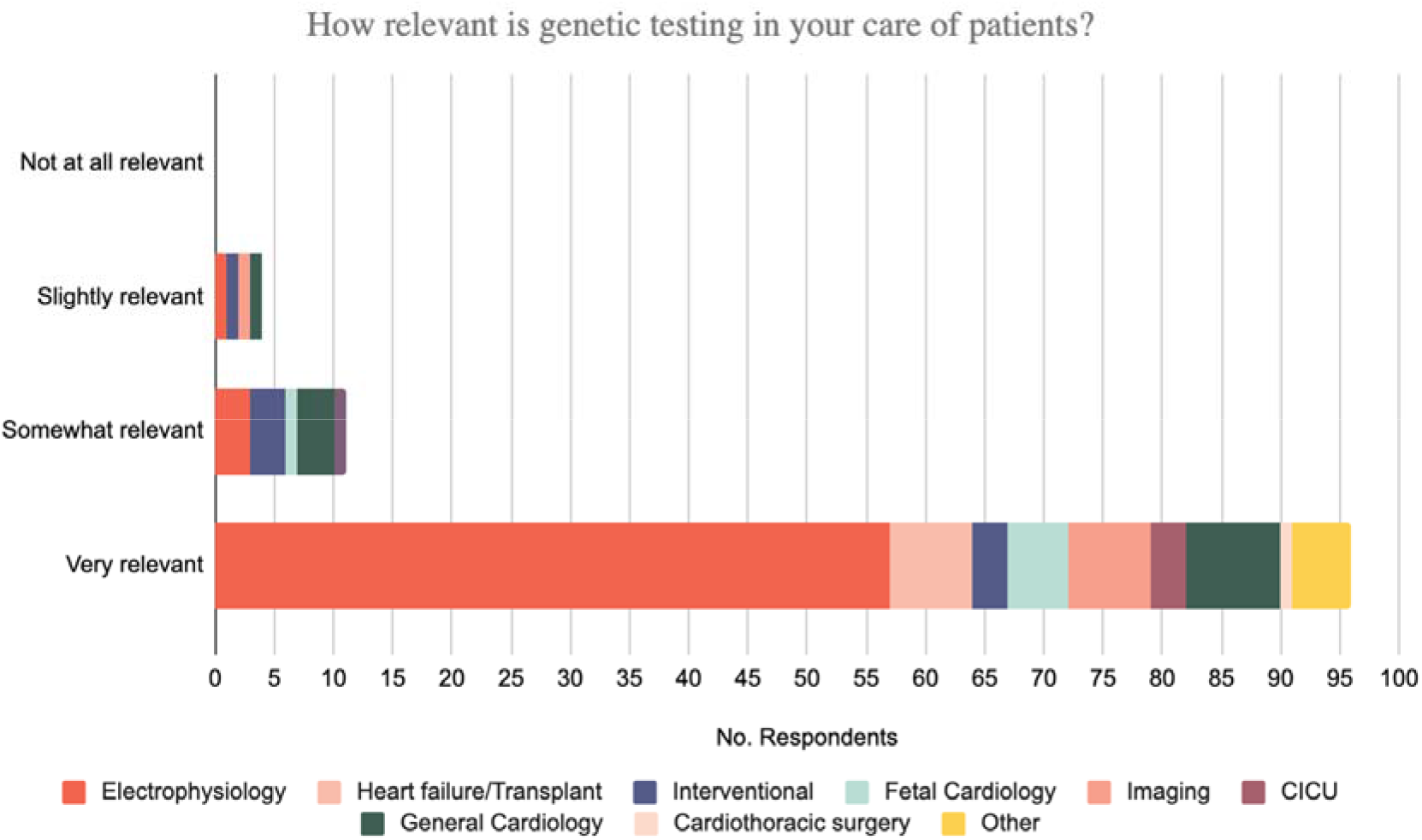
Provider Opinions on the Relevance of Genetic Testing *Responses to survey question 1 “How relevant is genetic testing in your care of patients” are delineated by provider specialty*.

The ordering trends of a few key specialists stood out and were investigated further: electrophysiologists, interventional cardiologists, and general cardiologists. These trends were groups or providers who reported either they consistently ordered genetic testing routinely or not at all. The findings of all providers and their reported use of genetic testing are depicted in Figure 2. Of electrophysiologists, ninety-six percent reported ordering genetic testing at least a few times per year at minimum. As expected, a high percentage, 80%, of electrophysiologists reported ordering genetic testing at least a few times per month. Interventional cardiologists were on the other end of the spectrum regarding ordering trends: fifty-seven percent of interventional cardiologists reported never ordering genetic testing or stated they did not know how often they order genetic testing. General cardiologists were somewhere in the middle with their ordering trends: seventy-five percent of general cardiologists reported ordering genetic testing a few times per year or never.

**FIGURE 2.**
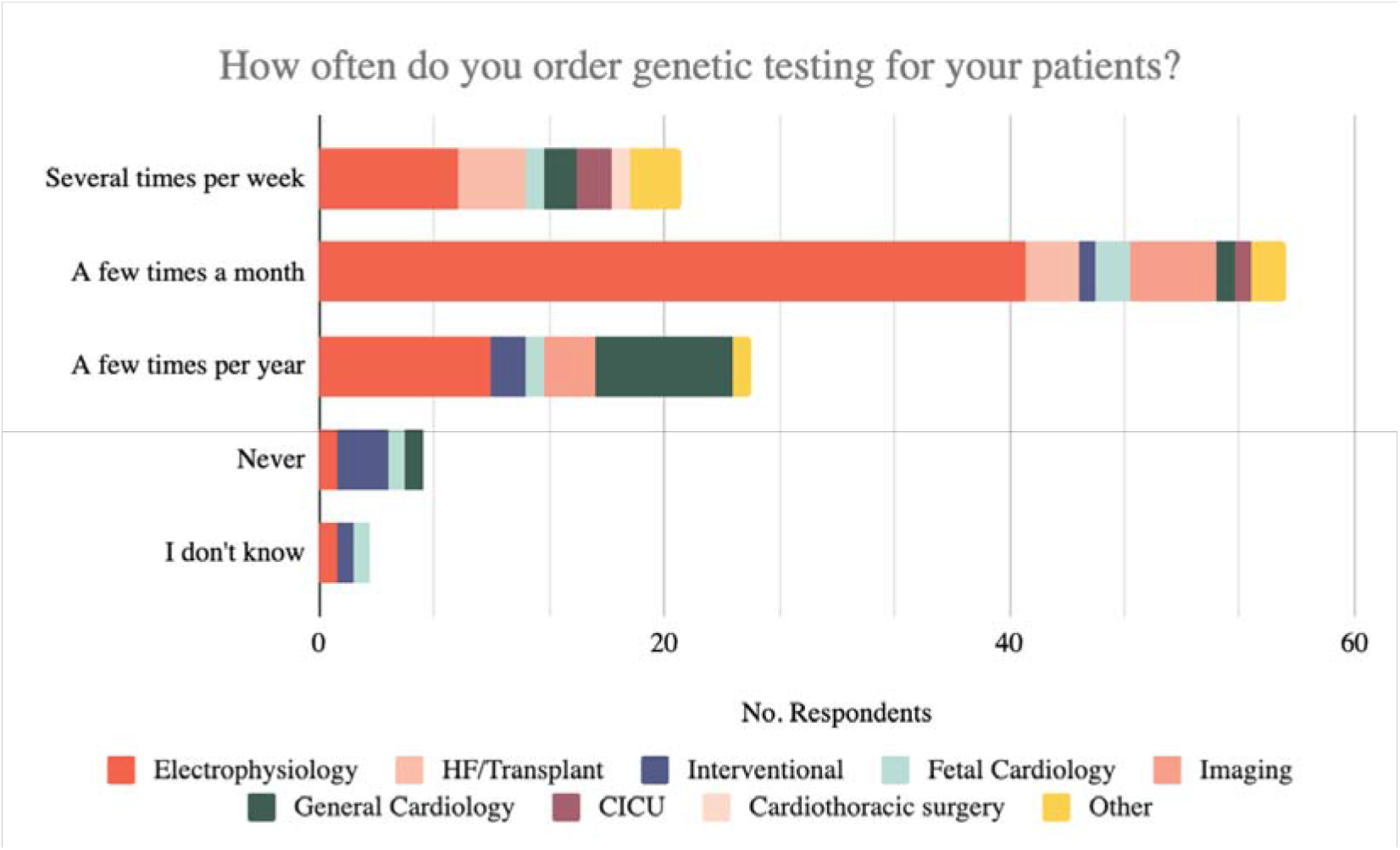
Provider Reported Use of Genetic Testing *Responses to survey question 11 “How often do you order genetic testing for your patients” are delineated by provider specialty*.

### Reported Barriers to Genetic Testing

Respondents were asked to report their comfort level with activities related to genetic testing including navigating insurance authorization and billing, interpreting genetic testing results for heritable cardiac conditions, and ability to communicate the results of genetic testing to patients. Thirty-five percent (n=39) of respondents reported being ‘uncomfortable’ with navigating insurance authorization and billing procedures. Based on respondents’ answers to this survey questions, we propose the ability to navigate insurance authorization and billing procedures is a potential barrier to the use of genetic testing by cardiovascular providers.

Genetic test interpretation and communication of genetic test results were not identified as being significant sources of provider discomfort. When respondents were asked to rank their comfort with interpreting genetic test results for heritable cardiac conditions based on previous training, thirty-six percent36% (n=40) stated they were ‘somewhat comfortable’. In response to a comfort level with communication of genetic testing results to the patient, thirty-eight percent38% (n=43) of respondents stated they were ‘somewhat comfortable’. These results are summarized in Figure 3.

**FIGURE 3.**
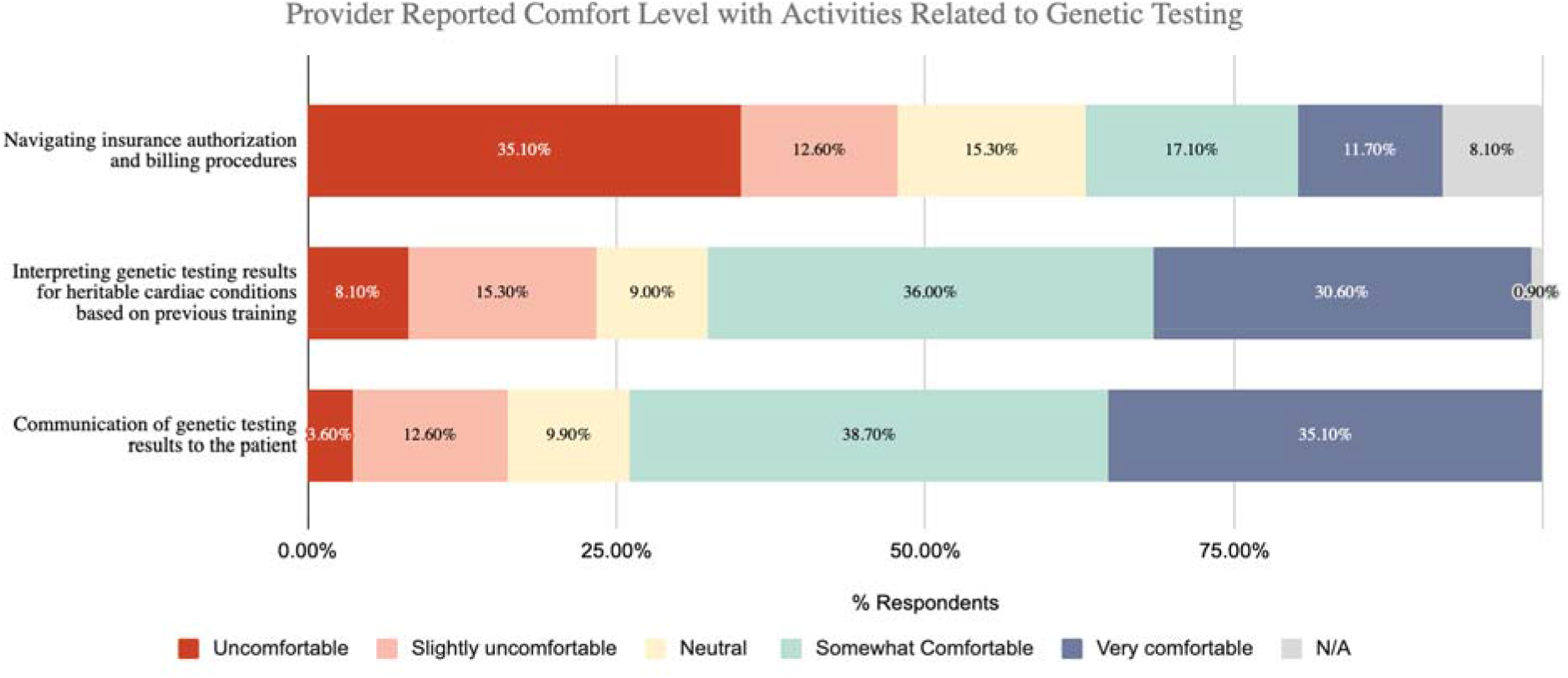
Reported Comfort Level with Activities Related to Genetic Testing *Responses to questions in section (4) of the survey regarding provider comfort with activities related to genetic testing are reported by percentage of respondents*.

### Influence of Provider Demographics on Genetic Testing

Statistical analyses were performed to determine if any particular demographic attributes of a provider influenced their reported opinion of, use of, or perceived barriers to the use of genetic testing. These findings are summarized in Table 2. Most significantly, we found that the provider’s specialty had an impact on how often they reported ordering genetic testing in practice as well as their reported comfort level with interpreting genetic test results for heritable cardiac conditions. In addition, we determined the provider’s geographic work setting had an impact on how often they reported ordering genetic testing in practice. Interestingly, provider geographic work setting also influenced opinion of genetic testing’s value for screening at-risk family members and for medication and device management.

**TABLE 2.**
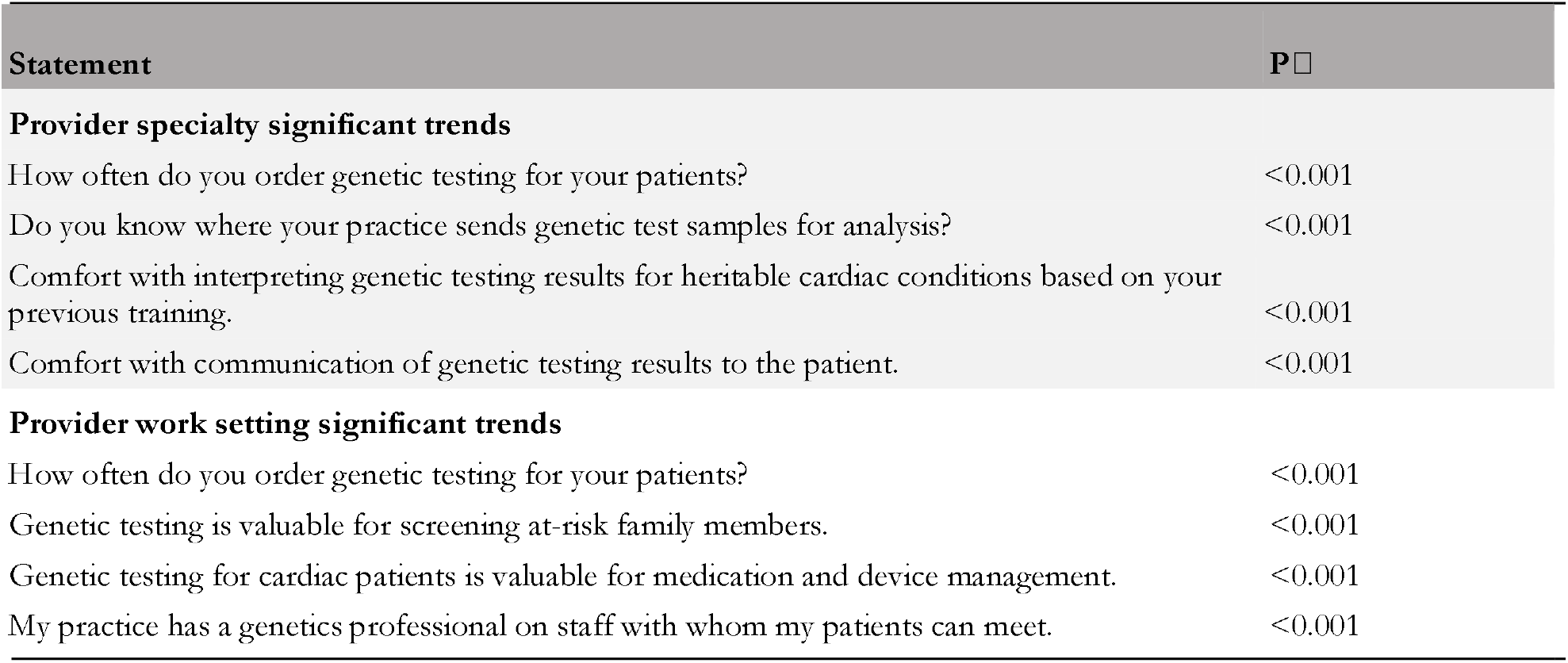
Provider Opinions on the Utility, Use, and Barriers of Genetic Testing in Practice. □: Chi-squared test with Bonferroni Correction (0.05/22; P<0.0022)

## DISCUSSION

Our study revealed that cardiovascular providers consider genetic testing relevant to their practice, which is important in understanding how to then implement it in practice. Guideline-directed medical management increasingly informs patient care. The guidelines from international organizations, such as the Heart Rhythm Society, have no specific legal territory and have no legally enforcing character. Nonetheless, they represent the state-of-the-art recommendations for patient diagnosis and management and set standards to which most practitioners aspire [20]. Our study tells us that the importance of genetic testing is understood by clinicians, but barriers exist to fully incorporating genetic testing into patient care. This study has practical applications regarding improvements to the uptake of genetic testing in cardiovascular medicine. Overall, the findings of this study suggest cardiovascular provider specialty impacts how often genetic testing is ordered in practice. The findings also suggest provider work setting impacts how often genetic testing is ordered in practice, provider opinion of the utility of genetic testing for family screening, and provider opinion on the utility of genetic testing for medication and device management. For example, in the case of potentially lethal and treatable conditions such as catecholaminergic polymorphic ventricular tachycardia (CPVT) or long QT syndrome (LQTS), it is the responsibility of the physician, preferably in conjunction with an expert genetics team, to communicate to the patient/family the critical importance of family screening, whether this be facilitated by cascade genetic testing or by broader clinical family screening [20]. Interestingly, the age range of patients a cardiovascular provider sees does not impact their opinion of, use of, or access to genetic testing. However, it should be noted that the sample size and queried population may have impacted this finding.

Based on the results of this study, navigating insurance authorization and billing procedures was an identified area of discomfort for cardiovascular providers. We suggest this as a targeted area for future cardiovascular provider education. This aligns with findings from other studies in the field [28, 32]. Designing educational materials that address the hurdles of insurance and billing is likely to be a useful strategy in addressing this area of concern for cardiovascular providers in the United States. Recommendations could be made for partnering with genetic counselors or local communities such as the SADS Foundation to help providers better understand insurance coverage. Automated modules about insurance or clear pathways of communication with genetic testing strategies could be useful in addressing this.

Interestingly, our study did not find significant provider discomfort in interpreting genetic testing. This could be due to the large majority of providers representing the specialty of electrophysiology. This group of providers has a longstanding history of utilizing genetic testing in patient care and understanding the underlying causes of inherited arrhythmias is an important component of clinical care. Other studies in the field [28, 32] have found provider discomfort with interpretation. Eighty percent of respondents (n=89) to this survey stated they would be interested in further training and education regarding genetic testing interpretation should their schedules allow, and 45% (n=50) stated they would be ‘likely’ to participate in an online webinar on genetic testing interpretation if prompted. Expanding the population size surveyed may reveal concerns about test interpretation, but from our evaluation, cardiologists specializing in electrophysiology as a group feel comfortable with genetic test results interpretation.

A limitation of this study was the limited sample size that was primarily comprised of electrophysiologists from urban university medical centers. This population is likely to have a self-reporting bias since we primarily contacted providers who associate with the SADS Foundation, a patient advocacy group that widely promotes genetic testing. The homogenous population of survey respondents limits the conclusions and applications that can be drawn from these results. It should also be noted that the survey developed and implemented in this study is not a validated tool and results from survey questions should be analyzed with this in mind.

## CONCLUSION

The need for genetic testing in cardiovascular medicine has been clearly defined and is part of the most recent expert consensus documents[20]. While respondents to this survey clearly acknowledge the relevance of genetic testing in the care of their patients, significant genetic testing care gaps for patients with heritable genetic testing conditions remain with few patients undergoing recommended genetic testing during their cardiac evaluation. This research captures certain provider characteristics-specialty and geographic work setting-that influence genetic testing ordering and uptake. Further research is needed to understand how to best address the disconnect between opinion and the use of genetic testing, and to continue improving the uptake of genetic testing for patients with inherited cardiac conditions.

## Supporting information

Supplemental Survey

## Data Availability

All data produced in the present study are available upon request to the authors.

## ACKNOWLEDGEMENTS

This manuscript is based on a research project conducted by Sierra Pond to fulfill the requirements of the University of Utah Master’s Degree in Genetic Counseling. This project was made possible by integral support from the Sudden Arrhythmia Death Syndromes (SADS) Foundation.

## REFERENCES

1. Walsh, R., et al., Evaluation of gene validity for CPVT and short QT syndrome in sudden arrhythmic death. Eur Heart J, 2022. 43(15): p. 1500–1510.

2. Priori, S.G. and C.A. Remme, Inherited conditions of arrhythmia: translating disease mechanisms to patient management. Cardiovasc Res, 2020. 116(9): p. 1539–1541.

3. Ingles, J., et al., Genetic Testing in Inherited Heart Diseases. Heart Lung Circ, 2020. 29(4): p. 505–511.

4. Stafford, F., et al., The role of genetic testing in diagnosis and care of inherited cardiac conditions in a specialised multidisciplinary clinic. Genome Med, 2022. 14(1): p. 145.

5. Spoonamore, K.G. and S.M. Ware, Genetic testing and genetic counseling in patients with sudden death risk due to heritable arrhythmias. Heart Rhythm, 2016. 13(3): p. 789–97.

6. Futema, M., et al., Genetic testing for familial hypercholesterolemia-past, present, and future. J Lipid Res, 2021. 62: p. 100139.

7. Knight, L.M., et al., Genetic testing and cascade screening in pediatric long QT syndrome and hypertrophic cardiomyopathy. Heart Rhythm, 2020. 17(1): p. 106–112.

8. Specterman, M.J. and E.R. Behr, Cardiogenetics: the role of genetic testing for inherited arrhythmia syndromes and sudden death. Heart, 2023. 109(6): p. 434–441.

9. Lioncino, M., et al., Hypertrophic Cardiomyopathy in RASopathies: Diagnosis, Clinical Characteristics, Prognostic Implications, and Management. Heart Fail Clin, 2022. 18(1): p. 19–29.

10. Harris, S.L. and M.E. Lindsay, Role of Clinical Genetic Testing in the Management of Aortopathies. Curr Cardiol Rep, 2021. 23(2): p. 10.

11. Kaufman, E.S., et al., Management of Congenital Long-QT Syndrome: Commentary From the Experts. Circ Arrhythm Electrophysiol, 2021. 14(7): p. e009726.

12. Brodie, O.T., Y. Michowitz, and B. Belhassen, Pharmacological Therapy in Brugada Syndrome. Arrhythm Electrophysiol Rev, 2018. 7(2): p. 135–142.

13. Black, R.M., et al., Projected impact of pharmacogenomic testing on medications beyond antiplatelet therapy in percutaneous coronary intervention patients. Pharmacogenomics, 2020. 21(7): p. 431–441.

14. Ingles, J., et al., Guidelines for genetic testing of inherited cardiac disorders. Heart Lung Circ, 2011. 20(11): p. 681–7.

15. Ackerman, M.J., et al., HRS/EHRA expert consensus statement on the state of genetic testing for the channelopathies and cardiomyopathies this document was developed as a partnership between the Heart Rhythm Society (HRS) and the European Heart Rhythm Association (EHRA). Heart Rhythm, 2011. 8(8): p. 1308–39.

16. Wilde, A.A.M., et al., European Heart Rhythm Association (EHRA)/Heart Rhythm Society (HRS)/Asia Pacific Heart Rhythm Society (APHRS)/Latin American Heart Rhythm Society (LAHRS) Expert Consensus Statement on the state of genetic testing for cardiac diseases. Europace, 2022. 24(8): p. 1307–1367.

17. Lui, D.T.W., A.C.H. Lee, and K.C.B. Tan, Management of Familial Hypercholesterolemia: Current Status and Future Perspectives. J Endocr Soc, 2021. 5(1): p. bvaa122.

18. Baumgartner, H., et al., 2020 ESC Guidelines for the management of adult congenital heart disease. Eur Heart J, 2021. 42(6): p. 563–645.

19. Writing Committee, M., et al., 2020 AHA/ACC guideline for the diagnosis and treatment of patients with hypertrophic cardiomyopathy: A report of the American College of Cardiology/American Heart Association Joint Committee on Clinical Practice Guidelines. J Thorac Cardiovasc Surg, 2021. 162(1): p. e23–e106.

20. Wilde, A.A.M., et al., European Heart Rhythm Association (EHRA)/Heart Rhythm Society (HRS)/Asia Pacific Heart Rhythm Society (APHRS)/Latin American Heart Rhythm Society (LAHRS) Expert Consensus Statement on the State of Genetic Testing for Cardiac Diseases. Heart Rhythm, 2022. 19(7): p. e1–e60.

21. Towbin, J.A., et al., 2019 HRS expert consensus statement on evaluation, risk stratification, and management of arrhythmogenic cardiomyopathy. Heart Rhythm, 2019. 16(11): p. e301–e372.

22. Murdock, D.R., et al., Genetic testing in ambulatory cardiology clinics reveals high rate of findings with clinical management implications. Genetics in Medicine, 2021. 23(12): p. 2404–2414.

23. Cohen, J.K., Most Patients With Inherited Heart Conditions Not Getting Guideline-Backed Genetic Testing. Precision Medicine Online, 2023.

24. Sturm, A.C., et al., Clinical Genetic Testing for Familial Hypercholesterolemia: JACC Scientific Expert Panel. J Am Coll Cardiol, 2018. 72(6): p. 662–680.

25. Longoni, M., et al., Real-world utilization of guideline-directed genetic testing in inherited cardiovascular diseases. Front Cardiovasc Med, 2023. 10: p. 1272433.

26. Dusic, E.J., et al., Barriers, interventions, and recommendations: Improving the genetic testing landscape. Front Digit Health, 2022. 4: p. 961128.

27. Smith, H.S., et al., Patient and Clinician Perceptions of Precision Cardiology Care: Findings From the HeartCare Study. Circ Genom Precis Med, 2022. 15(6): p. e003605.

28. Lopez Santibanez Jacome, L., et al., Practitioners’ Confidence and Desires for Education in Cardiovascular and Sudden Cardiac Death Genetics. J Am Heart Assoc, 2022. 11(7): p. e023763.

29. Aatre, R.D. and S.M. Day, Psychological issues in genetic testing for inherited cardiovascular diseases. Circ Cardiovasc Genet, 2011. 4(1): p. 81–90.

30. Harris, P.A., et al., Research electronic data capture (REDCap)--a metadata-driven methodology and workflow process for providing translational research informatics support. J Biomed Inform, 2009. 42(2): p. 377–81.

31. Harris, P.A., et al., The REDCap consortium: Building an international community of software platform partners. J Biomed Inform, 2019. 95: p. 103208.

32. Spoonamore, K.G. and N.M. Johnson, Who Pays? Coverage Challenges for Cardiovascular Genetic Testing in U.S. Patients. Front Cardiovasc Med, 2016. 3: p. 14.

