## Supplemental Survey for "Identifying barriers to genetic testing in subspecialty cardiac care"

**Survey**

*The purpose of this study is to determine the attitudes of cardiovascular providers and their use of genetic testing in the care of their patients. Participation should take approximately ten minutes. Your participation in this survey is voluntary. You may refuse to take part in the research and exit the survey at any time without penalty.*
*Survey data will be secure and only accessible to members of the research team as well as the University of Utah Institutional Review Board (IRB) if requested. No identifying personal information will be collected and therefore all responses will remain completely anonymous.*
*The information will be collected and stored in a survey software program called REDCap and used for statistical analysis by the research team. Survey responses may be incorporated into a scientific publication.*
*If you have any questions please contact the lead investigator, Sierra Pond, at. Contact the IRB if you have questions regarding your rights as a research participant. You may contact the IRB if you have questions, complaints, or concerns which you do not feel you can discuss your concerns with the study investigator. The University of Utah IRB may be reached at 801-581-3803 or by email at.*
*By participating in the survey, you are giving your consent to participate in this research. Thank you for your willingness to participate.*

**Do you consent to participate in this research study?**
**-Yes**
**-No (If you select no, the survey will close)**

**Inclusion Criteria**

A) Are you a licensed MD, DO, PA, RN, or APRN in the United States? (*Lopez Santibanez Jacome et al, 2022)
   a. Yes
   b. No

B) If yes, do you provide direct patient care in the United States? (*Lopez Santibanez Jacome et al, 2022)
   a. Yes
   b. No
   If no, the survey closes.

C) Are you currently in training in your education?
   a. Yes
      i. If yes, please select the stage of training: resident/fellow/student/other
   b. No

D) What best describes your area of specialty within cardiology? (*Lopez Santibanez Jacome et al, 2022)
   a. Electrophysiology
   b. Heart failure/Transplant
   c. Interventional
   d. Fetal Cardiology
   e. Imaging
   f. Pulmonary Hypertension

g. Dyslipidemia
h. CICU
i. General Cardiology
j. Cardiothoracic surgery
k. Other, not listed

**PART I: Genetic Testing Utility**

*This group of ten questions will examine how you regard genetic testing in the context of patient care.*

1. How relevant is genetic testing in your care of patients?
    a. Not at all relevant
    b. Slightly relevant
    c. Somewhat relevant
    d. Very relevant

Please indicate your opinion on the following statements.

| | Strongly agree | Somewhat agree | Neither agree nor disagree | Somewhat disagree | Disagree |
|---|---|---|---|---|---|
| 2. **Genetic testing is valuable for cardiac patients in establishing a diagnosis.** | ○ | ○ | ○ | ○ | ○ |
| 3. **Genetic testing aids in ruling out certain conditions on the differential.** | ○ | ○ | ○ | ○ | ○ |
| 4. **Genetic testing is valuable for screening at-risk family members.** | ○ | ○ | ○ | ○ | ○ |
| 5. **Genetic testing for cardiac patients is valuable for anticipating patient prognosis.** | ○ | ○ | ○ | ○ | ○ |
| 6. **Genetic testing is valuable for patient management.** | ○ | ○ | ○ | ○ | ○ |
| 7. **Genetic testing for cardiac** | ○ | ○ | ○ | ○ | ○ |

**patients is valuable for medication and device management.**

8. In your ideal patient care setting, when would you incorporate genetic testing?
    a. Never
    b. Pre-diagnosis
    c. At the time of clinical diagnosis
    d. Post clinical diagnosis
    e. Other, please explain

**PART II: Implementing Genetic Testing in Practice**

*This group of four questions will ask about how/when/if you implement genetic testing in the care of cardiovascular patients.*

9. About how many patients in your practice do you think are eligible for genetic testing?
    a. None, I do not see the utility of genetic testing in my practice.
    b. 1-10%
    c. 11-25%
    d. 26-50%
    e. >50%
    f. I don't know

10. How often do you order genetic testing for your patients?
    a. Several times per week
    b. A few times a month
    c. A few times per year
    d. Never

11. Do you know where your practice sends genetic test samples for analysis?
    a. Yes
    b. No
    c. Some of the time
    d. Not applicable, I do not see the utility of genetic testing in my practice.

12. My practice has the following: select all that apply (multi-select radio buttons)

| | | | | |
|---|---|---|---|---|
| A genetics professional on staff with whom my patients can meet | Contact with an external genetics professional to whom I can refer my patients | Contact with a laboratory genetics professional | No contact with a genetics professional | Other, please explain |

**PART III: Barriers to Genetic Testing in Practice**
*This group of seven questions will ask if you experience any barriers in implementing genetic testing in the care of cardiovascular patients.*

Please indicate your comfort level with the following activities related to genetic testing:

| | Very comfortable | Somewhat comfortable | Neither comfortable nor uncomfort-able | Slightly uncomfor-table | Uncomfor-table |
|---|---|---|---|---|---|
| 13. **Navigating insurance authorization and billing procedures:** | ○ | ○ | ○ | ○ | ○ |
| 14. **Interpreting genetic testing results for inheritable cardiac conditions based on your previous training:** | ○ | ○ | ○ | ○ | ○ |
| 15. **Communication of genetic testing results to the patient:** | ○ | ○ | ○ | ○ | ○ |

16. Does time in your clinic schedule factor into whether you feel you can discuss genetic testing with a patient?
    a. Yes
    b. No
    c. I do not see the utility of discussing genetic testing with my patients.

17. Did your residency/fellowship/clinical rotations include training on genetic testing?
    a. Yes
    b. No

18. Are you interested in further training/education regarding genetic testing interpretation?
    a. Yes
    b. If my schedule allows
    c. No

19. If an online webinar on genetic testing interpretation was offered to you, how likely would you be to participate?
    a. Very likely
    b. Likely
    c. Unlikely
    d. Very unlikely

20. What resources are you aware of and/or utilize when you get back a genetic test result you are uncertain of how to interpret? (select all that apply)
    a. Varsome
    b. Franklin
    c. gnomAD
    d. ClinVar
    e. ClinGen
    f. USCS Genome Browser
    g. Other, please state.

**PART IV: Demographics**
*These final questions will ask about specific demographic elements of your medical career.*

21. In what state do you practice?
    Drop down menu

22. What age range of patients do you see?
    a. Pediatrics
    b. Adult
    c. Both pediatric and adult
    d. Post-mortem
    e. Laboratory/diagnostic

23. How long have you been practicing medicine? (*Lopez Santibanez Jacome et al, 2022)
    a. Still in training
    b. < 5 years
    c. 5-10 years
    d. 10-15 years
    e. >15 years

24. Which of the following best describes your primary work setting? (*Lopez Santibanez Jacome et al, 2022)
    a. Private Group Practice
    b. Private Solo Practice
    c. University Medical Center
    d. Private Hospital/Medical Facility
    e. Public Hospital/Medical Facility
    f. Diagnostic Laboratory/Testing Laboratory
    g. Other: please specify

25. In which geographic region is your primary work setting? (*Lopez Santibanez Jacome et al, 2022)
    a. Urban
    b. Suburban
    c. Rural
